# A Longitudinal Analysis of COVID-19 Lockdown Stringency on Sleep and Resting Heart Rate Measures across 20 Countries

**DOI:** 10.1101/2021.03.15.21253668

**Authors:** Ju Lynn Ong, TeYang Lau, Mari Karsikas, Hannu Kinnunen, Michael WL Chee

## Abstract

Lockdowns imposed to stem the spread of COVID-19 massively disrupted the daily routines of many worldwide, but studies to date are mostly confined to observations within a limited number of countries, based on subjective reports and survey from specific time periods during the pandemic. We investigated associations between lockdown stringency and objective sleep and resting-heart rate measures in 113,000 users of a consumer sleep tracker across 20 countries from Jan-Jul 2020. With stricter lockdown measures, midsleep times were universally delayed, particularly on weekdays, while midsleep variability and resting heart rate declined. These shifts (midsleep: +0.09 to +0.58 hours; midsleep variability: –0.12 to –0.26 hours; resting heart rate: –0.35 to –2.08 bpm) correlated with the severity of lockdown across different countries and highlight the graded influence of mobility restriction and social isolation on human physiology.

## Introduction

Sans social obligations, sleep-wake timings are determined by the interaction between an individual’s circadian clock and the timing of natural light exposure ^1,2^. However, in most industrialized societies, organized work with its complex web of values and activities have insidiously and incrementally transformed our natural sleep patterns. Modern-day workers are likely to go to bed later, sleep less regularly and get exposed to greater stress than their predecessors ^3-5^.

Recent lockdowns imposed around the world to contain the spread of COVID-19 resulted in massive disruption of daily routines surrounding work and face-to-face social interactions ^6-8^. In theory, closure of workplaces and schools as well as sports, entertainment and social hubs should free up much time for neglected uses of time, including sleep. Work-from-home arrangements can afford individuals more latitude to adopt their preferred sleep-wake timings ^9^. In turn, this could reduce variability in sleep timing that, when high, has been linked to poor sleep quality, impaired health and well-being as well as metabolic abnormalities ^10-14^. On the other hand, being in a state of lockdown might cause anxiety and depression for many, reducing the amount of sleep obtained ^15^. In addition, reduced morning light exposure and increased evening light exposure from screens could lead to progressive delays in chronotype ^16^ and a loss of circadian rhythm entrainment ^17^. Perhaps as a consequence of the aforesaid factors that push sleep in opposite directions, reports of sleep behavior during the lockdown suggest only modest gains in sleep time ^15,18-23^ and are mixed depending on region/demographic surveyed.

To date, studies documenting COVID-19 pandemic changes in sleep behavior and their potential health implications have largely been confined to observations within a country or a limited number of countries ^6-8,18,20,22,24-32^. As prevailing social norms and the severity of lockdowns differ across countries, the generalizability of such reports is unclear. Additionally, most studies have utilized questionnaires in which baseline data was inferred from participant recollection past sleep habits. Finally, few studies have followed the temporal evolution of sleep through both lockdown and the lifting of restrictions.

To fill these gaps in our understanding of sleep behavior throughout this challenging period, we analyzed nocturnal sleep behavior and resting heart rate of ∼113,000 users of the Oura ring sleep tracker from Jan – Jul 2020, using an equivalent period in 2019 as a baseline control. The Oura ring is a novel multisensor device that uses motion, heart-rate and temperature sensors to detect sleep/wake states (https://ouraring.com/). It has been validated in various population groups with sleep-wake detection performance comparable to that of research grade actigraphy and polysomnography ^33-37^. This tracker allowed for large streams of longitudinal data to be collected during this period with minimal user effort, enabling analysis from a prepandemic baseline period, through lockdowns and lifting of restrictions in the 20 countries, spanning regions in North America, Europe, Asia and Oceania.

We sought to (a) assess how two critical sleep parameters were influenced by pandemic-related lockdown and subsequent partial lifting of restrictions, (b) determine the extent to which these sleep parameters were influenced by the severity of lockdown measures and (c) relate how alterations in sleep behaviour affected resting heart rate, an indicator of cardiovascular health. To optimize clarity and utility of sleep data from a public health perspective, we focused on average mid-sleep time and sleep variability. The former provides a compact measure that is influenced by later bed and wake times linked to academic and health outcomes ^38,39^, while the latter is an indicator of irregular sleep/wake patterns - also associated with negative health and cognitive outcomes independently of sleep duration ^10,12,40,41^. Lockdown stringency was assessed using scores extracted from the Oxford Government Response Tracker ^42^, which considers factors such as school and workplace closures, cancellations of public events and gatherings and closures of public transport.

## Results

### Regional and Global Trends in Sleep and Resting Heart Rate

Data was obtained from randomly selected Oura users for two equivalent periods in 2019 and 2020. Sample size, mean age and mean BMI values by country of included users are detailed in Table S1. Only countries with a minimum of 500 users were included in subsequent analyses.

Midsleep time as well as resting heart rate showed weekday-weekend differences whereby weekends were associated with later midsleep time as well as higher resting heart rate (Figure 1). Comparing data collected on the same day in 2020 and 2019, it is evident that when lockdowns were most severe across 20 countries, midsleep time shifted later, midsleep variability decreased and resting heart rate decreased. Inspection of sleep duration trends in 2020 also indicated that most countries showed increases during the months of Mar-May 2020 compared to Jan 2020 (ie, before lockdowns occurred; Figure S1A). Later weekday midsleep times were contributed by delayed bedtimes and commensurately more delayed wake times (elevated troughs of orange relative to blue midsleep time courses in Figure S1 B-C), reduced the weekday-weekend difference in midsleep times, in turn reducing midsleep variability.

**Figure 1.**
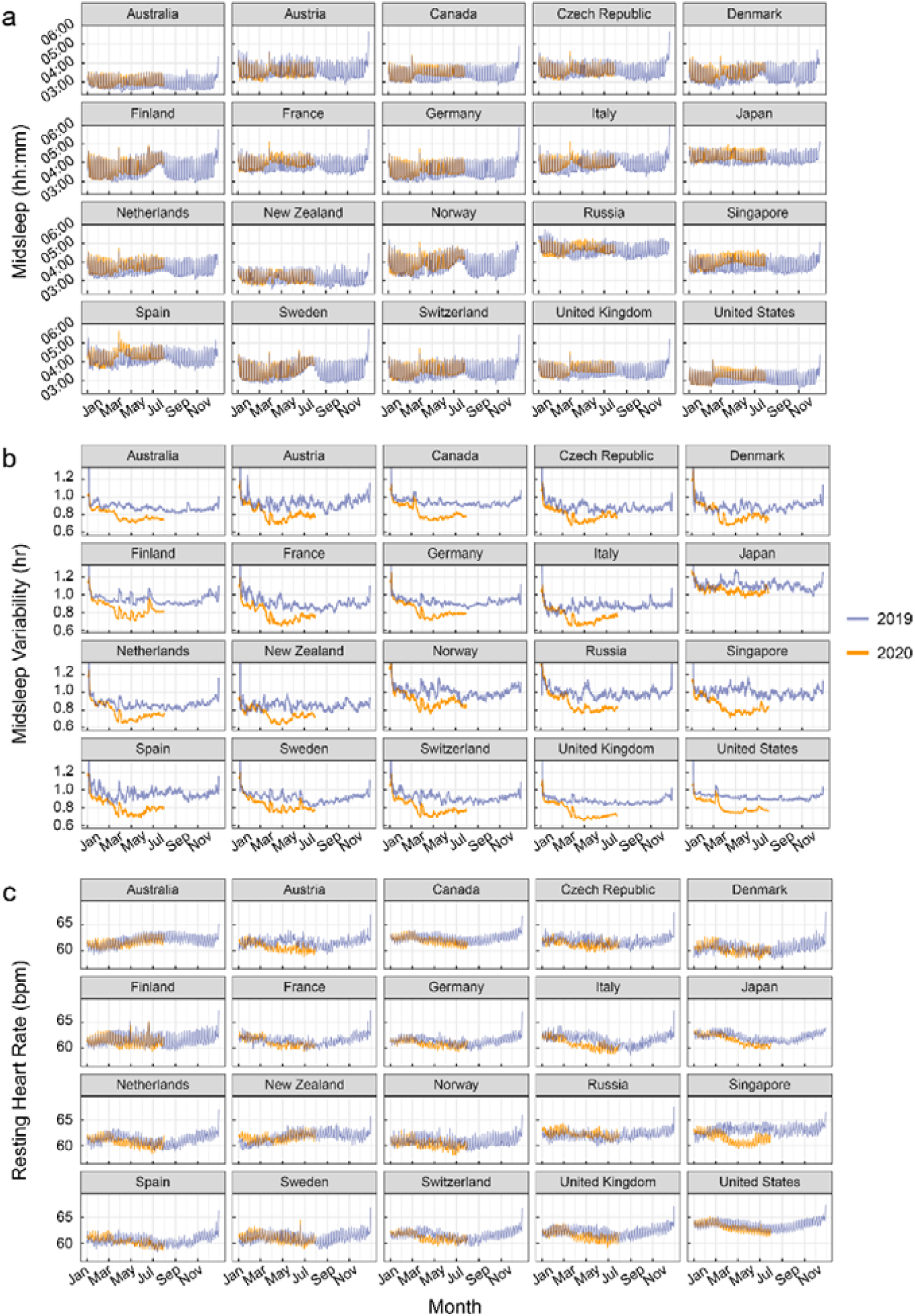
Sleep patterns and resting heart rate measures by country. **(a)** Midsleep time, (**b)** Midsleep variability and (**c)** Resting heart rate from Jan to Jul 2020 (orange curves), compared to Jan-Dec 2019 (purple curves). Dates in 2019 were shifted in order to ensure a matching by day of the week. Daylight savings time (social clocks shifted later by 1h) began in March in select countries of the Northern Hemisphere, and ended in April in Australia and New Zealand (social clocks shifted earlier by 1h), explaining sudden shifts in midsleep time and variability on these dates.

### Lockdown Stringency Modulates Changes in Sleep and Resting Heart Rate

Mirroring the heterogeneity in the severity of lockdowns, the shifts in midsleep time and sleep regularity differed widely across countries as evidenced by the high heterogeneity *I*^2^ statistic (>75%). Forest plots (Figure 2) depict country-level, month-by-month trends in the lockdown related shifts in midsleep time, midsleep variability and resting heart rate obtained by comparing monthly averages of data from comparable days in 2020 and 2019. Pooled effects derived from a random effects meta-analyses representing global changes by month are shown below each forest plot. In general, the largest shifts in midsleep time (+0.09 to +0.58 hours), midsleep variability (–0.12 to –0.26 hours) and resting heart rate (–0.35 to –2.08 bpm) occurred in April and May show when most countries imposed their strictest lockdown measures. Conversely the gradually lifting of restrictions from around June was accompanied by a return to patterns recorded in the previous year.

**Figure 2.**
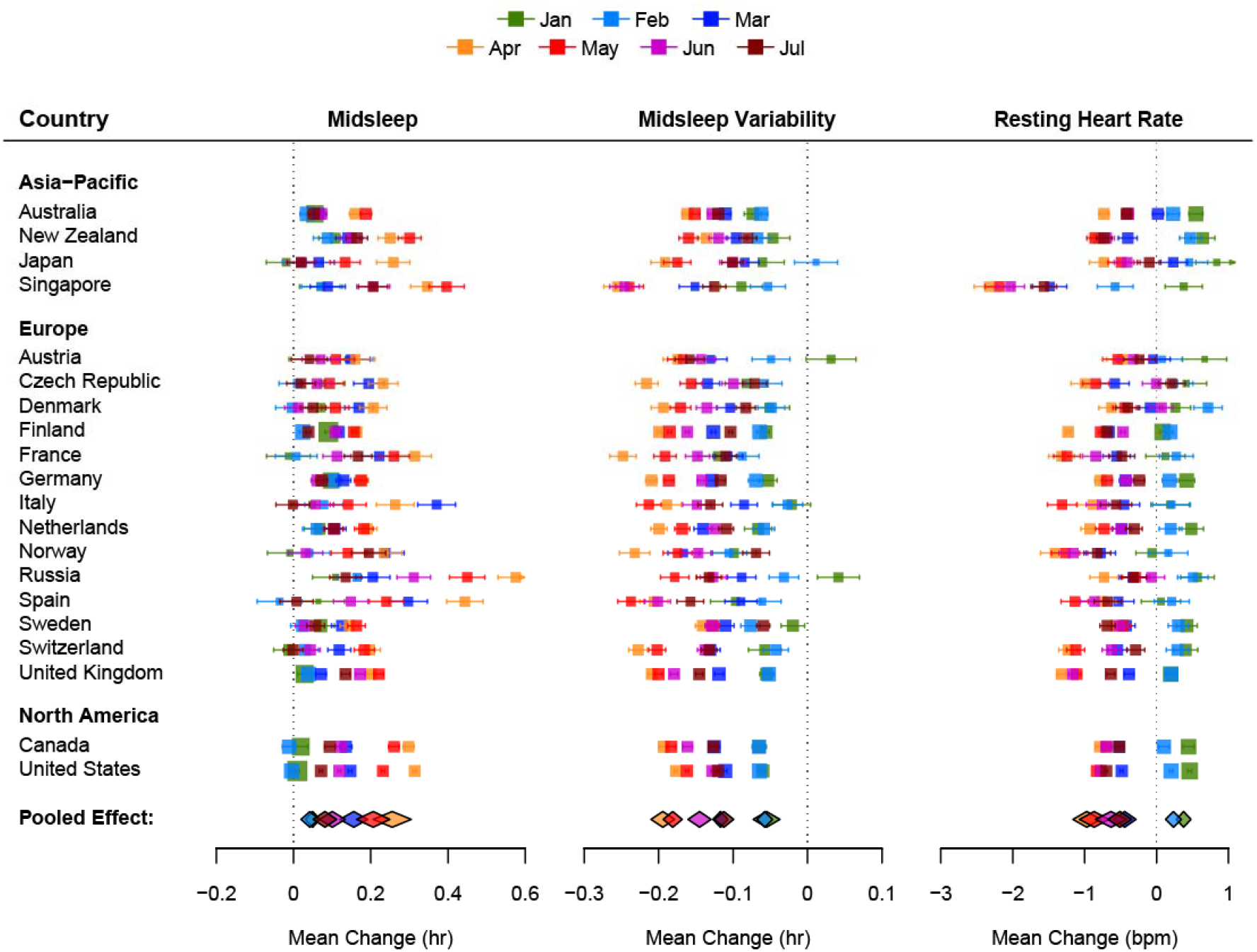
Forest plots of the average change in midsleep time, midsleep variability and resting heart rate by month and country. The size of the colored squares is proportional to the sample size of each country while whiskers indicate the mean and 95% confidence interval of the estimated difference between 2020 and 2019 by month (Jan-Jul). The overall pooled effect across countries for each month is represented by the colored diamonds below each plot.

Multilevel modelling revealed that the extent to which sleep timings and resting heart rate shifted during the pandemic’s most pressing early months, could be largely explained by the severity of the stringency index (Figure 3 and Tables S2-S4).

**Figure 3.**
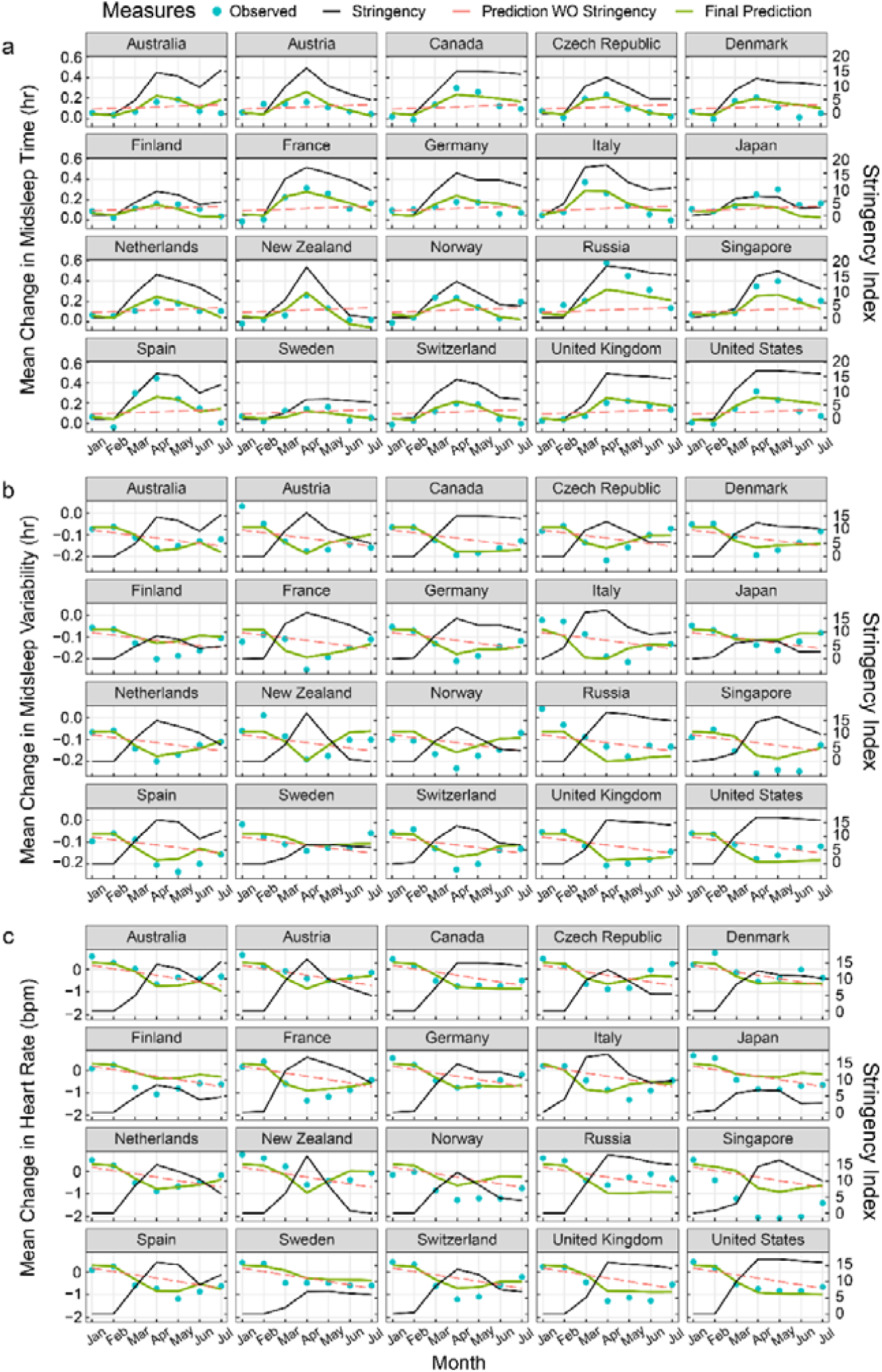
Prediction of changes in sleep patterns and resting heart rate by lockdown stringency. Plots for fitted models (green curves) across 20 countries estimating changes in (**a)** Midsleep Time, (**b)** Midsleep Variability and (**c)** Resting heart rate using lockdown stringency as a predictor. Observed mean differences between 2020 and 2019 (blue dots), lockdown stringency (black line) and predictions without lockdown stringency in the model (red dashed line) are also shown for comparison.

Marginal *R*^2^ values increased from 0.02, 0.18 and 0.28 in baseline models to 0.56, 0.60 and 0.57 for midsleep time, midsleep variability and resting heart rate respectively when stringency index was included a predictor in the model. For each unit increase in stringency index, midsleep time was delayed by 0.96 min, midsleep variability decreased by 0.46 min and resting heart rate decreased by 0.06 bpm.

### Changes in Sleep Patterns Predict Changes in Resting Heart Rate

Finally, in models predicting 2020-2019 changes in resting heart rate associated with changes in midsleep time, midsleep variability and absolute sleep duration, we found that while each of the three variables significantly predicted resting heart rate in separate models, the model fit was significantly improved when all three predictors were included in the model (marginal *R*^2^ = 0.75, Table 1). In this model (Model 5), midsleep variability was also shown to be the strongest predictor of resting heart rate, wherein an hour increase in the standard deviation of midsleep variability predicted a 5.12 increase in bpm, while an hour increase in midsleep time only predicted a 1.25 decrease in bpm. Sleep duration in 2020 also no longer significantly predicted resting heart rate changes in Model 5 (Table 1). The final model fit using Model 5 was fitted to each country (Figure 4). It was evident that the fitted curves mirrored the trends of changes in midsleep variability observed in each country.

**Table 1.**
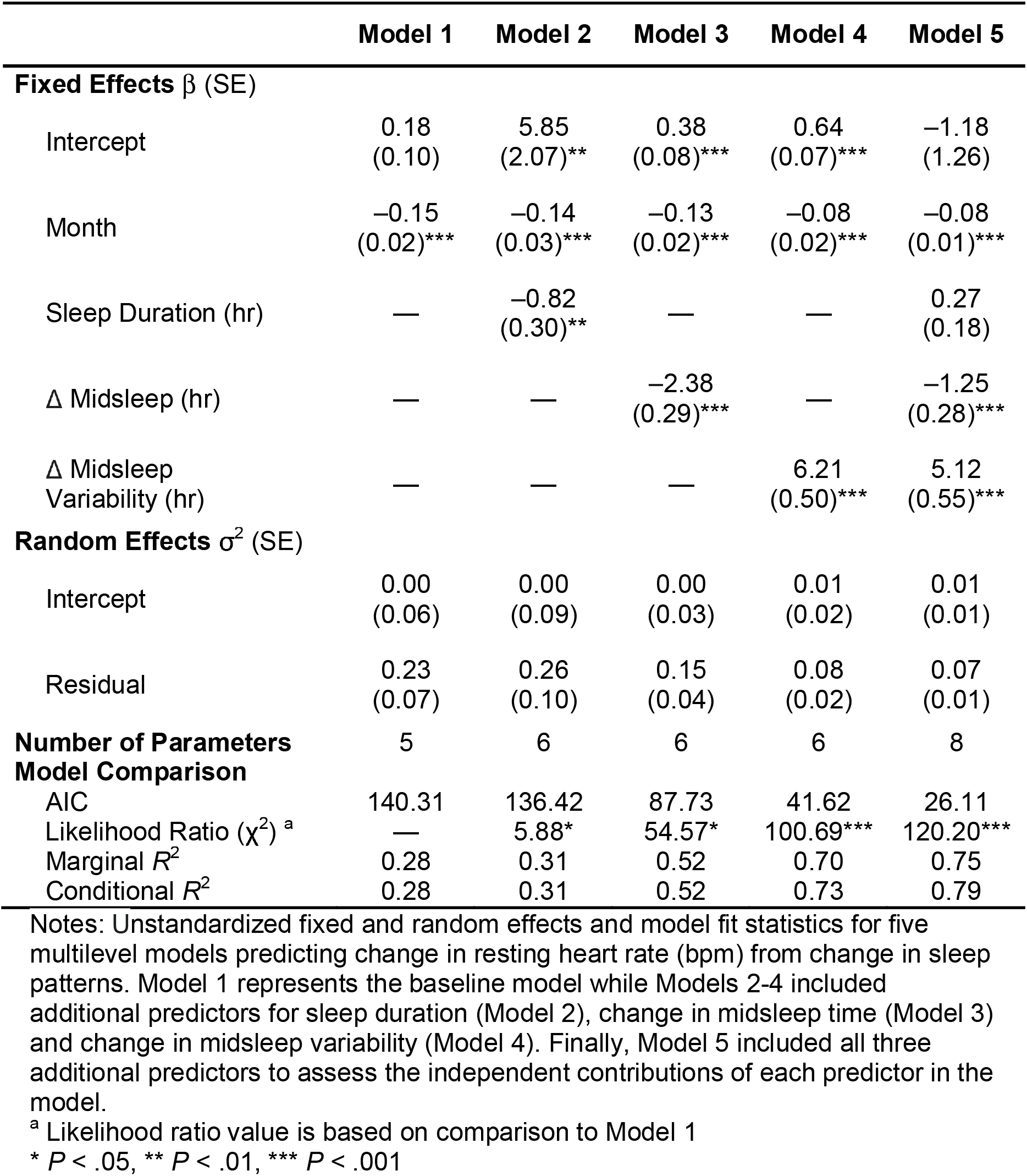
Model fit statistics predicting changes in resting heart rate.

|  | **Model 1** | **Model 2** | **Model 3** | **Model 4** | **Model 5** |
| --- | --- | --- | --- | --- | --- |
| **Fixed Effects $\beta$ (SE)** |  |  |  |  |  |
| Intercept | 0.18<br>(0.10) | 5.85<br>(2.07)** | 0.38<br>(0.08)*** | 0.64<br>(0.07)*** | -1.18<br>(1.26) |
| Month | -0.15<br>(0.02)*** | -0.14<br>(0.03)*** | -0.13<br>(0.02)*** | -0.08<br>(0.02)*** | -0.08<br>(0.01)*** |
| Sleep Duration (hr) | — | -0.82<br>(0.30)** | — | — | 0.27<br>(0.18) |
| $\Delta$ Midsleep (hr) | — | — | -2.38<br>(0.29)*** | — | -1.25<br>(0.28)*** |
| $\Delta$ Midsleep<br>Variability (hr) | — | — | — | 6.21<br>(0.50)*** | 5.12<br>(0.55)*** |
| **Random Effects $\sigma^2$ (SE)** |  |  |  |  |  |
| Intercept | 0.00<br>(0.06) | 0.00<br>(0.09) | 0.00<br>(0.03) | 0.01<br>(0.02) | 0.01<br>(0.01) |
| Residual | 0.23<br>(0.07) | 0.26<br>(0.10) | 0.15<br>(0.04) | 0.08<br>(0.02) | 0.07<br>(0.01) |
| **Number of Parameters** | 5 | 6 | 6 | 6 | 8 |
| **Model Comparison** |  |  |  |  |  |
| AIC | 140.31 | 136.42 | 87.73 | 41.62 | 26.11 |
| Likelihood Ratio ($\chi^2$)<sup>a</sup> | — | 5.88* | 54.57* | 100.69*** | 120.20*** |
| Marginal $R^2$ | 0.28 | 0.31 | 0.52 | 0.70 | 0.75 |
| Conditional $R^2$ | 0.28 | 0.31 | 0.52 | 0.73 | 0.79 |

Notes: Unstandardized fixed and random effects and model fit statistics for five multilevel models predicting change in resting heart rate (bpm) from change in sleep patterns. Model 1 represents the baseline model while Models 2-4 included additional predictors for sleep duration (Model 2), change in midsleep time (Model 3) and change in midsleep variability (Model 4). Finally, Model 5 included all three additional predictors to assess the independent contributions of each predictor in the model.
<sup>a</sup> Likelihood ratio value is based on comparison to Model 1
\* $P < .05$ , \*\* $P < .01$ , \*\*\* $P < .001$

**Figure 4.**
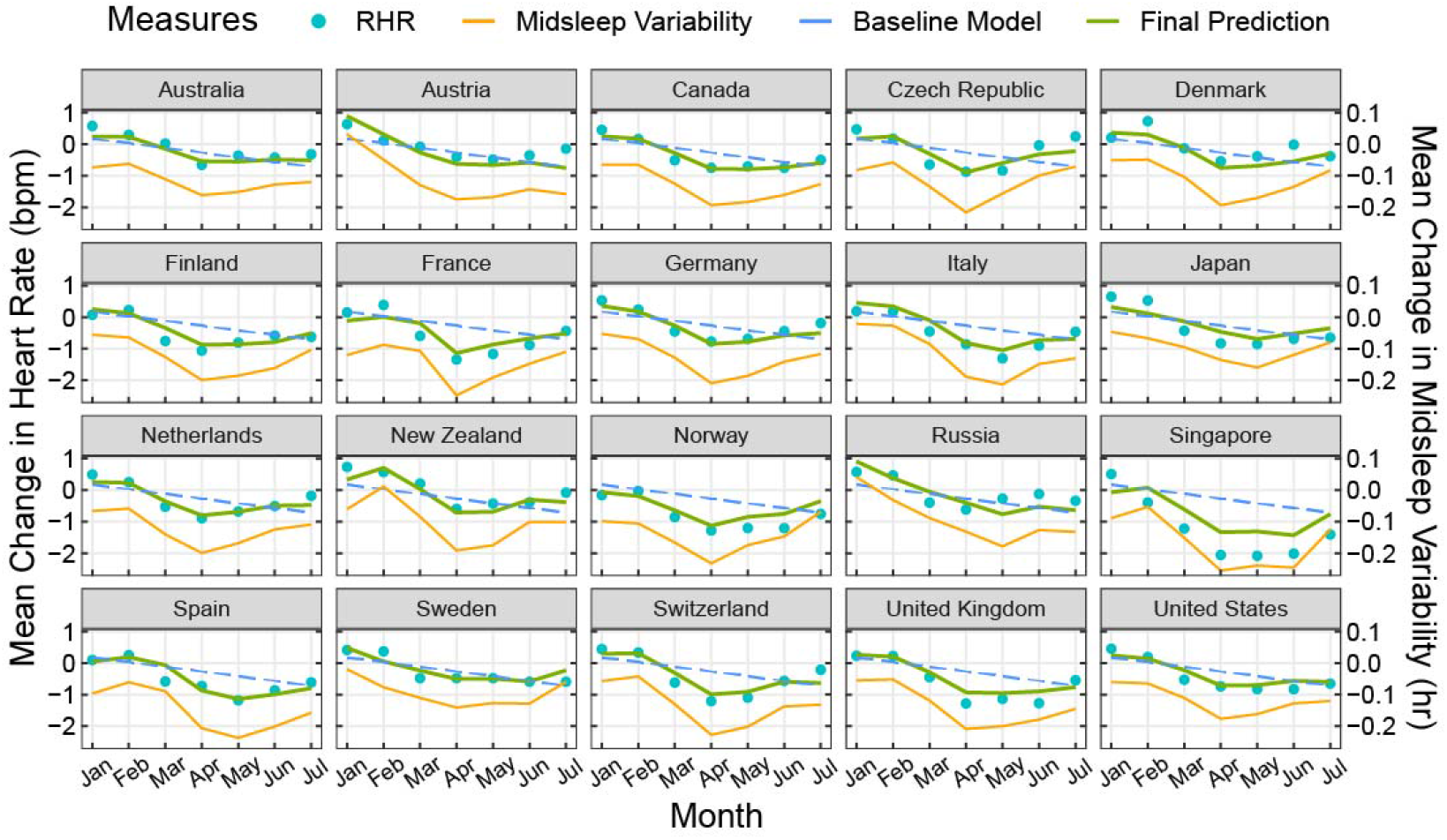
Prediction of changes in resting heart rate by changes in sleep patterns. Plots showing final fitted models (green curves) across 20 countries predicting changes in resting heart rate (2020-2019) using changes in midsleep time, changes in midsleep variability, and sleep duration in 2020 as time-varying predictors in the same model. Observed mean differences between 2020 and 2019 (blue dots) and predictions without sleep variability in the model (blue dashed line) are also shown for comparison. Sleep variability was found to be the strongest predictor of resting heart rate and is shown here in orange solid line for visualization purposes.

## Discussion

In all 20 countries across 4 continents, objective data obtained over successive years showed that pandemic-related lockdowns delayed sleep midpoint, reduced sleep variability and reduced resting heart rate as a function of the prevailing lockdown severity. Favorable resting heart rate decreases were most strongly related to reduction in sleep variability even after accounting for sleep duration. Although these changes were generalized across geographies and cultures, they unwound with easing of lockdowns.

Later timing of sleep has been associated with higher rates of metabolic dysfunction and cardiovascular disease ^43-46^. while deviations from normal bedtimes have been shown to elevate resting heart rate ^47^. There is also a risk that absence or attenuation of social cues together with reduced morning light exposure and increased evening light exposure from screens could lead to a loss of circadian rhythm entrainment ^17^. However, freed from the usual obligations to wake up earlier than preferred during lockdown and removal of commuting time, the negative effects of later bedtimes were partially made up for by later wake times and reduced weekday-weekend sleep timing differences, with resultant gains in sleep regularity – results similarly echoed in prior work ^18,20,22^. The latter can have positive effects on cardiometabolic health ^40^ possibly reflected here in reduced resting heart rate. While a lower resting heart rate is most often associated with higher physical fitness, studies examining physical activity following lockdown have found declines of up to 40% ^48,49^, making improved fitness an unlikely contributor in this context. Instead, our data indicated that at the country-level, sleep variability alone explained 70% of the variance in resting heart rate.

Both sleep variability and resting heart rate tended to drop most in countries where the strictest lockdown measures were implemented (e.g. Singapore). Conversely, Sweden had relatively lenient lockdown measures, and the impact on sleep and resting heart rate changes were smaller. On a month-to-month level, the time courses of resting heart rate were closely correlated with those of midsleep timing/variability across multiple countries. Of particular interest, the lifting of restrictions was accompanied by a rise in midsleep variability accompanied by a corresponding uptick in resting heart rate.

Our analysis also critically compared equivalent days across successive years. Raw measures in individual countries contain weekday-weekend, holiday and seasonality effects (e.g. in 2019) whereby weekends and holidays are associated with later sleep midpoints and higher resting heart rates. Conversely, summer is associated with later sleep midpoints, but with reduced sleep variability and lower resting heart rate. Seasonality effects are modulated by residential latitude and are phase-opposite in the Southern Hemisphere (Australia, New Zealand) compared to the Northern Hemisphere, while being practically absent in equatorial countries like Singapore ^50^. Seasonality effects were in fact, modulated during the lockdown, with an apparent advance in the appearance of summertime in the Northern hemisphere and apparent prolongation of summertime in the South.

The benefits of improving sleep on health have economic impact and are increasingly recognized ^51^. A large, time-use study suggested that interventions to increase sleep should concentrate on delaying morning start time for work and educational activities, increasing sleep opportunities and reducing commute times ^52^. Creating greater opportunity for sleep by working from home and giving workers some flexibility in sleeping according to preferred schedules ^53^ could yield benefits to both productivity and sleep health if properly implemented ^9,54^. To realize sustained improvements in sleep behavior, a critical area to address is the erosion of boundaries between work and home life, and a growing expectation for workers to be ‘always on’ and reachable using information and communications technology ^55,56^. For example, in France, there are laws governing the ‘right to disconnect’, to protect employees from having to engage in work-related electronic communications beyond working hours ^56^.

The explosive expansion of video-conferencing have made functional home-based work and learning widely accessible – something unlikely had the current pandemic occurred just two decades ago. The precipitous growth in remote work and learning was reflected in a 10%-60% rise in internet traffic within OECD countries during the early lockdown period in Mar-Apr 2020 ^57^. The National Bureau of Economic Research found that for over 3 million users across 16 metropolitan areas, COVID-19 related mobility restrictions lengthened workdays by 48.5 min from the sending of emails outside regular working hours ^58^. One network service provider reported an increase of 1-3 work hours per day in the US, UK, France, Spain, Canada and the Netherlands. In lieu to time saved from not having to commute, some started work earlier but ended at their habitual time ^59^. Outside work, online gaming platforms and social media activity also increased ^60^. Facebook reported increases of 100% on voice over internet calls and 50% in text messages on WhatsApp, Facebook Messenger and Instagram platforms during lockdown. For those with young children, adapting to home-based learning may also have displaced work time later. Some workers could also deliberately procrastinate sleep to regain a sense of control of personal time use. Together, these new daily routines could have contributed to progressively delayed midsleep timing. Future studies, supplemented by tools like ecological momentary assessments, would do well to understand heterogeneity in individual reactions to the blurring of work/non-work boundaries merits, and to include a wider demographic, particularly shift workers and lower income persons who live outside urban centers.

While this study highlights strengths of being able to rapidly and remotely assess the impact of various intervention policies on sleep and resting heart rate, there are a few limitations to consider. (1) Only sleep periods between 4-12 hours were analyzed. Shorter sleep periods could increase in frequency with work from home arrangements, but is challenging to detect and distinguish from other brief periods of sedentary behavior, e.g. sitting in bed reading a book or watching television. (2) Oura users typically come from middle to upper class households who could be more cushioned by the impact of COVID-19 and have flexible work arrangements. (3) Sleep quality measures were not obtained, which could be affected by anxiety from the impending loss of jobs or contracting the disease. However, one study showed unchanged or even improved sleep quality during the lockdown, particularly once shift workers and individuals who showed symptoms of COVID-19 were excluded from the analyses ^5^. (4) As these data were extracted from a large wearable database, we were not able to obtain information about occupations, shift work status, free vs. work days or caregiving responsibilities of these users. Those with additional childcare responsibilities due to school closures or have had a member of the household fall ill might also have had to work late hours in order to catch up on work. Not-withstanding these limitations, our model based on stringency indices was able to capture >50% of the variance in sleep and resting heart rate measures, indicating that it is a key predictor of sleep and resting heart rate trends during this period.

In sum, the use of large-scale wearable data revealed consistent and geographically widespread nature of the correlation between lockdown severity and shift to delayed but more regular sleep with reductions in resting heart rate measures. These findings should spur governments to consider the secondary health impact of various policies and interventions during this period and beyond.

## Methods

### Dataset

Data from 2019 was used as the reference year, representing a typical annual cycle and seasonal variation, for example, in terms of holidays, amount of light/daylength, and incidence of influenza-like illnesses. In order to ensure alignment by day of the week between 2019 and 2020, days were shifted before further computations were conducted. Due to the extra leap day in 2020, data was shifted by one day in Jan and Feb 2020 and by two days in Mar-Jul. Local timestamps for each country and time zone were also utilized, which included shifts that reflect daylight savings start (Mar-Apr in the Northern Hemisphere, Sep-Oct in the Southern Hemisphere) and end points (Apr in the Southern Hemisphere, Sep-Nov in the Northern Hemisphere). Paired differences between matching days in 2019 and 2020 for users who had valid data in both timepoints were then computed and included in subsequent analyses.

Each valid sleep period was defined as the longest sleep episode for each day, with time in bed between 4-12 hours. Three major variables were then extracted for each of these sleep periods: **(1) Midsleep time** was computed as the midpoint between bedtime and wake time, representing a proxy for circadian phase/chronotype ^61^ (2) **Midsleep variability** was computed using a rolling 7-day standard deviation of midsleep times, representing a proxy for sleep regularity, and (3) **Resting heart rate** which was computed as an average of 5-min heart rate measures during the sleep period. Average resting heart rates <30 bpm and >100bpm were removed as these were likely to represent physiologic or device anomalies. Due to an algorithm update in the spring of 2019 that affected computation of sleep duration by delaying bedtimes and advancing wake times, this could not be compared between years, however, absolute sleep duration in 2020 was included as an additional variable in time-varying models predicting resting heart rate.

Age and BMI information was self-reported by users upon app registration, and entered into models as potential covariates. This study was exempt from review by the National University of Singapore Institutional Review Board, as analysis involved the use of datasets stored without identifiers.

### Computation of Stringency Index

Publicly available measures of restriction severity were extracted from the Oxford COVID-19 Government Response Tracker,^42^ focusing on 7 subscales believed to be most reflective of movement controls. These scales consisted of (1) school closures [0-3], (2) workplace closures [0-3], cancellation of public events [0-2], restrictions on public gatherings [0-4], closures of public transport [0-2], stay-at-home requirements [0-3] and restrictions on internal movements [0-2]. These 7 subscales were summed up into a single stringency index [range: 0-19] and a mean value was computed for every month from January to July for each country.

### Quantification of Regional and Global Trends in Sleep and Resting Heart Rate

Changes in sleep and resting heart rate measures were derived for each month within each country separately, by first computing differences between equivalent days in Jan–Jul 2019 and 2020, and then averaging these differences by month. To estimate global (pooled) changes, separate random-effects meta-analyses by month were conducted for each predictor of interest – midsleep time, midsleep variability (standard deviation of the midsleep time over a 7-day rolling window) and resting heart rate. Meta-analyses were conducted using the R package ‘metafor’ ^62^. As there was evidence of high statistical heterogeneity between country estimates by month (Cochrane’s Q; *P* < .05, *I*^2^ > 75%), pooled estimates were weighted by the inverse variance of estimators for each country plus the estimated variance between countries.

### Quantification of the Effect of Lockdown Stringency on Changes in Sleep and Resting Heart Rate

In order to quantify the effect of lockdown stringency on the heterogenous changes in sleep and resting heart rate patterns across countries, we ran multilevel growth curve models (MLMs) based on a sequential model-building approach. Multilevel models account for correlations between months within each country by allowing each country to have its own intercept. A null or baseline model is first constructed, and subsequent models consisting of the baseline model + additional explanatory variables were added sequentially to assess if the more complex model improved the overall model fit using a likelihood ratio test with degrees of freedom equal to the number of extra parameters. A significant likelihood ratio test indicates that the extra parameters improved the fit of the model to the data.

For each of the variables of interest (midsleep time, midsleep variability, resting heart rate), baseline MLMs (Model 1) were first estimated using country as a random intercept, month as a fixed effect, and a first-order autoregressive term. The latter was included to account for the nature of correlated time points in the repeated variable (month). Finally, in Model 2, we included the average lockdown stringency index by month as a time-varying factor to Model 1, in order to examine the overall effect of lockdown stringency on sleep and resting heart rate measures.

### Quantification of the Effect of Changes to Sleep Patterns on Changes to Resting Heart Rate

To test our hypothesis that changes to sleep patterns (midsleep time and midsleep variability) would lead to associated changes in resting heart rate, we conducted further MLM analyses with changes in resting heart rate as the dependent variable and changes in midsleep time and changes in midsleep variability as explanatory variables. Sleep duration in 2020 was also included as an additional variable of interest in this model. A baseline MLM (Model 1) with a random intercept, month as fixed effect, and first-order autoregressive structure was first constructed. Age and BMI were entered as covariates, but were subsequently removed as they did not significantly improve the baseline model. Next, in Models 2-4, sleep duration in 2020, changes to midsleep time and changes to midsleep variability were added as time-varying predictors in separate models. Finally, in Model 5, all three sleep measures were entered in at the same time to assess the independent contributions of each predictor in the model.

All MLMs were estimated using the full information maximum likelihood method and performed using the nlme package in R (version 3.6.1). Marginal and conditional R^2^ values for mixed models are calculated based on ^63^. Notably, the marginal R^2^ only takes into account the variance of the fixed effects, while the conditional R^2^ takes both fixed and random effects into account.

## Data Availability

Aggregate data are available from the authors upon reasonable request.

## Contributions

All authors contributed to study design, interpretation of the reported results and drafting/revision of the manuscript. M.K. and H.K. contributed to data extraction efforts from the Oura database. J.L.O., T.L. and M.K. contributed to data analysis and production of figures and tables.

## Data Availability

Aggregate data are available from the authors upon reasonable request.

## Abbreviations

MLM: Multilevel Model

