## Supplementary Materials for "A Longitudinal Analysis of COVID-19 Lockdown Stringency on Sleep and Resting Heart Rate Measures across 20 Countries"

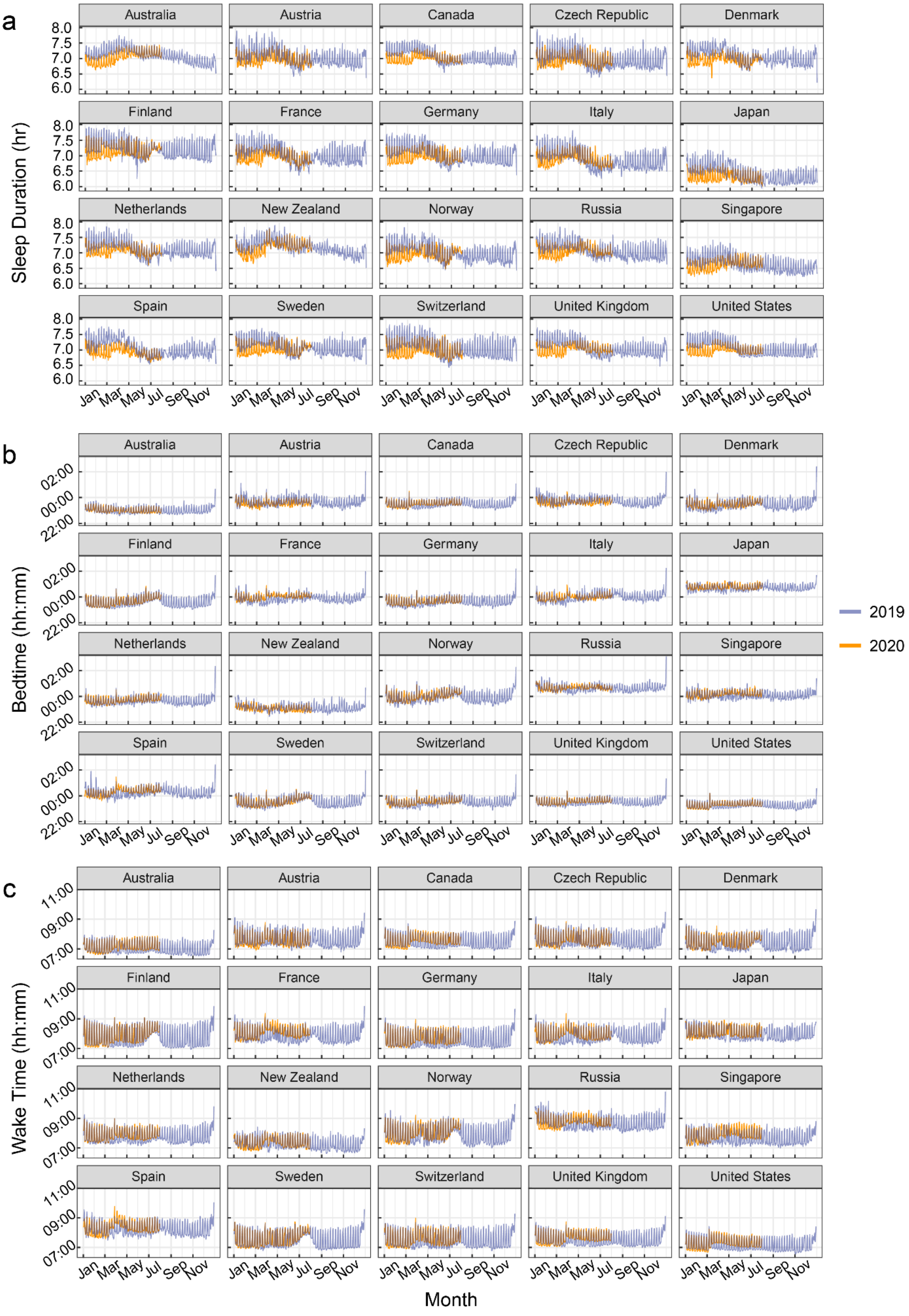

**Figure S1. Sleep duration, bed and wake times by country.**
**(a)** Sleep Duration, (**b)** Bedtimes and (**c)** Wake times in 2020 (orange curves) compared to Jan-Dec 2019 (purple curves). An algorithm change that delayed bedtimes and advanced wake times occurred in the Spring of 2019, which affect direct comparisons between 2020 and 2019 - observable by a downward shift in estimates for sleep duration in Jan 2020 compared to Jan 2019 (before lockdowns occurred). However, as there was no algorithm change in 2020, estimates during the year itself can still be reliably compared.

**Table S1. Number of subjects contributing to this study, their mean age and mean BMI by country.**

| Country | N* | Age | | BMI | |
|---|---|---|---|---|---|
| | | mean | std | mean | std |
| Australia | ≥5,000 | 41.40 | 11.32 | 25.26 | 4.09 |
| Europe Austria | 1,000 | 39.71 | 11.37 | 24.67 | 3.84 |
| Canada | ≥5,000 | 42.39 | 11.81 | 25.29 | 4.33 |
| Czech Republic | 1,000 | 36.83 | 8.90 | 25.49 | 5.07 |
| Denmark | 1,000 | 41.13 | 11.31 | 24.90 | 4.33 |
| Finland | ≥10,000 | 43.34 | 11.08 | 25.94 | 4.32 |
| France | 1,000 | 41.28 | 12.17 | 23.64 | 3.48 |
| Germany | 4,000 | 41.06 | 11.23 | 24.60 | 3.83 |
| Italy | 1,000 | 41.93 | 11.95 | 24.23 | 3.74 |
| Japan | 2,000 | 35.42 | 10.02 | 22.43 | 3.42 |
| Netherlands | 1,000 | 41.15 | 11.38 | 24.23 | 3.73 |
| New Zealand | 1,000 | 43.80 | 11.48 | 25.40 | 4.12 |
| Norway | 1,000 | 39.04 | 11.27 | 24.36 | 3.33 |
| Russia | 1,000 | 36.41 | 8.64 | 23.51 | 3.71 |
| Singapore | 1,000 | 40.97 | 9.00 | 24.02 | 3.88 |
| Spain | 1,000 | 42.37 | 11.67 | 23.86 | 3.48 |
| Sweden | 2,000 | 42.04 | 11.46 | 24.49 | 3.72 |
| Switzerland | 1,000 | 42.98 | 11.58 | 24.09 | 3.77 |
| United Kingdom | ≥5,000 | 42.90 | 11.06 | 25.05 | 4.35 |
| United States | ≥10,000 | 45.02 | 12.45 | 25.40 | 4.59 |

Notes: * Rounded to the closest 1000

**Table S2. Unstandardized fixed and random effects and model fit statistics for two multilevel models of midsleep time (min) – without and with lockdown stringency.**

| | **Model 1** | **Model 2** |
|---|---|---|
| **Fixed Effects** β (SE) | | |
| Intercept | 5.58 (1.23)*** | 3.65 (0.87)*** |
| Month | 0.43 (0.31) | -1.11 (0.24)*** |
| Stringency Index | — | 0.96 (0.08)*** |
| **Random Effects** $\sigma^2$ (SE) | | |
| Intercept | 0.00 (6.29) | 1.93 (2.87) |
| Residual | 37.84 (8.36) | 16.4 (3.37) |
| **Number of Parameters** | 5 | 6 |
| **Model Comparison** | | |
| AIC | 883.88 | 782.58 |
| Likelihood Ratio ($\chi^2$) [a] | — | 103.30*** |
| Marginal $R^2$ | 0.02 | 0.56 |
| Conditional $R^2$ | 0.02 | 0.61 |

Notes: * p < 0.05, ** p < 0.01, *** p < 0.001
[a] Likelihood ratio value is based on comparison to Model 1

**Table S3. Unstandardized fixed and random effects and model fit statistics for two multilevel models of midsleep variability (min) – without and with lockdown stringency.**

| | **Model 1** | **Model 2** |
|---|---|---|
| **Fixed Effects** β (SE) | | |
| Intercept | -4.68 (0.62)*** | -3.75 (0.47) |
| Month | -0.73 (0.16)*** | 0.01 (0.13) |
| Stringency Index | — | -0.46 (0.04)*** |
| **Random Effects** $\sigma^2$ (SE) | | |
| Intercept | 0.00 (1.65) | 0.00 (0.93) |
| Residual | 9.58 (2.17) | 5.09 (1.21) |
| **Number of Parameters** | 5 | 6 |
| **Model Comparison** | | |
| AIC | 689.55 | 599.17 |
| Likelihood Ratio ($\chi^2$) [a] | — | 92.38*** |
| Marginal $R^2$ | 0.18 | 0.60 |
| Conditional $R^2$ | 0.18 | 0.60 |

Notes: * p < 0.05, ** p < 0.01, *** p < 0.001
[a] Likelihood ratio value is based on comparison to Model 1

**Table S4. Unstandardized fixed and random effects and model fit statistics for two multilevel models of resting heart rate (bpm) – without and with lockdown stringency.**

| | **Model 1** | **Model 2** |
|---|---|---|
| **Fixed Effects** β (SE) | | |
| Intercept | 0.18 (0.10) | 0.30 (0.08)*** |
| Month | -0.15 (0.02)*** | -0.05 (0.02)* |
| Stringency Index | — | -0.06 (0.01)*** |
| **Random Effects** $\sigma^2$ (SE) | | |
| Intercept | 0.00 (0.06) | 0.00 (0.05) |
| Residual | 0.23 (0.07) | 0.15 (0.06) |
| **Number of Parameters** | 5 | 6 |
| **Model Comparison** | | |
| AIC | 140.31 | 57.83 |
| Likelihood Ratio ($\chi^2$) [a] | — | 84.47*** |
| Marginal $R^2$ | 0.28 | 0.57 |
| Conditional $R^2$ | 0.28 | 0.57 |

Notes: * p < 0.05, ** p < 0.01, *** p < 0.001

[a] Likelihood ratio value is based on comparison to Model 1
